# Distinct explanations underlie gene-environment interactions in the UK Biobank

**DOI:** 10.1101/2023.09.22.23295969

**Authors:** Arun Durvasula, Alkes L. Price

## Abstract

The role of gene-environment (GxE) interaction in disease and complex trait architectures is widely hypothesized, but currently unknown. Here, we apply three statistical approaches to quantify and distinguish three different types of GxE interaction for a given trait and E variable. First, we detect locus-specific GxE interaction by testing for genetic correlation (*r_g_*) < 1 across E bins. Second, we detect genome-wide effects of the E variable on genetic variance by leveraging polygenic risk scores (PRS) to test for significant PRSxE in a regression of phenotypes on PRS, E, and PRSxE, together with differences in SNP-heritability across E bins. Third, we detect genome-wide proportional amplification of genetic and environmental effects as a function of the E variable by testing for significant PRSxE with no differences in SNP-heritability across E bins. Simulations show that these approaches achieve high sensitivity and specificity in distinguishing these three GxE scenarios. We applied our framework to 33 UK Biobank traits (25 quantitative traits and 8 diseases; average *N*=325K) and 10 E variables spanning lifestyle, diet, and other environmental exposures. First, we identified 19 trait-E pairs with *r_g_* significantly < 1 (FDR<5%) (average *r_g_*=0.95); for example, white blood cell count had *r_g_*=0.95 (s.e. 0.01) between smokers and non-smokers. Second, we identified 28 trait-E pairs with significant PRSxE and significant SNP-heritability differences across E bins; for example, BMI had a significant PRSxE for physical activity (P=4.6e-5) with 5% larger SNP-heritability in the largest versus smallest quintiles of physical activity (P=7e-4). Third, we identified 15 trait-E pairs with significant PRSxE with no SNP-heritability differences across E bins; for example, waist-hip ratio adjusted for BMI had a significant PRSxE effect for time spent watching television (P=5e-3) with no SNP-heritability differences. Across the three scenarios, 8 of the trait-E pairs involved disease traits, whose interpretation is complicated by scale effects. Analyses using biological sex as the E variable produced additional significant findings in each of the three scenarios. Overall, we infer a significant contribution of GxE and GxSex effects to complex trait and disease variance.

## Introduction

Although gene-environment (GxE) interactions have long been thought to impact the genetic architecture of diseases and complex traits^1–4^, the overall contribution of these effects remains unclear. Previous studies have detected GxE at a limited number of specific loci^5–7^ (including studies that associated genotype to phenotypic variance without knowing the underlying E variable^8–12)^. Previous studies have also proposed variance components methods for detecting genome-wide contributions of GxE to complex trait heritability^13–18^, but these methods have not been applied at biobank scale across a broad range of traits. Thus, the overall contribution of GxE to trait architectures is currently unknown. In addition, the relative importance of different types of GxE (e.g., locus-specific GxE, genome-wide effects of E on genetic variance, genome-wide effects of E on both genetic and environmental variance) is currently unclear. Studies of GxSex interaction face similar challenges^19–24^.

Here, we apply three statistical approaches to quantify and distinguish three different types of GxE interaction for a given trait and E variable. First, we detect locus-specific GxE interaction by testing for genetic correlation^25^ (*r_g_*) < 1 across E bins. Second, we detect genome-wide effects of the E variable on genetic variance by leveraging polygenic risk scores^26,27^ (PRS) to test for significant PRSxE^28,29^ in a regression of phenotypes on PRS, E, and PRSxE, together with differences in SNP-heritability^30–34^ across E bins. Third, we detect genome-wide proportional amplification of genetic and environmental effects as a function of the E variable by testing for significant PRSxE with no differences in SNP-heritability across E bins. We analyze 33 traits from the UK Biobank^35^ (25 quantitative traits and 8 diseases; average *N*=325K), quantifying the contributions of each type of GxE effect across 10 E variables spanning lifestyle, diet, and other environmental exposures, as well as contributions of GxSex effects.

## Results

### Overview of methods

We aim to detect genome-wide GxE, i.e., GxE effects aggregated across the genome. We consider three potential scenarios that give rise to genome-wide GxE for a given trait and E variable (**Figure 1a**). In the first scenario (Imperfect genetic correlation), there is an imperfect genetic correlation across E bins due to different SNP effect sizes in different E bins. In the second scenario (Varying genetic variance), there are differences in SNP-heritability across E bins due to uniform amplification of SNP effect sizes across E bins; the environmental variance may either remain constant or vary across E bins. In the third scenario (proportional amplification), the genetic and environmental variance vary proportionately across E bins due to proportionate scaling of SNP effect sizes and environmental effect sizes across E bins, so that SNP-heritability remains the same across E bins. We conceptualize these three scenarios as acting at different levels in a hierarchy that leads from genetic variants to pathways to complex traits or disease (see **Discussion**).

**Figure 1.**
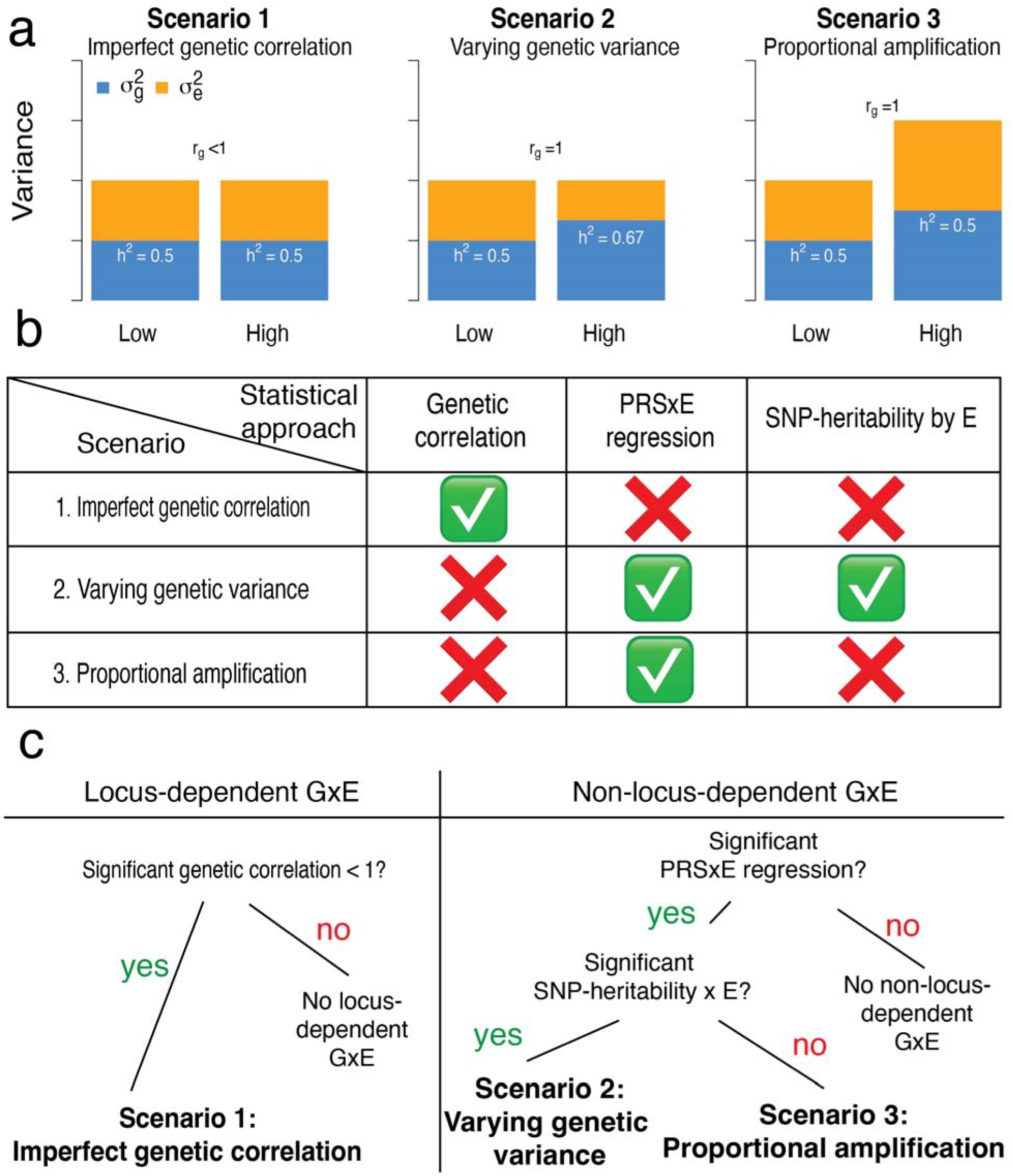
Overview of 3 GxE Scenarios and statistical approaches to detect and distinguish between them. (a) Relative values of genetic (blue) and environmental (orange) variance in each Scenario. (b) Statistical approaches to detect and distinguish between each Scenario. (c) Flow chart for classifying results into Scenarios.

The three scenarios can be formalized under the following model:

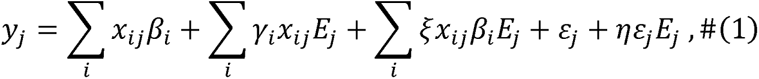

where *y_i_* denotes the phenotype for individual *j, x_ij_* denotes the genotype of individual *j* at SNP *i*, *β_i_* denotes the effect size of SNP *i γ_i_* denotes SNP-specific GxE effects, *E_j_* denotes the E variable value for individual *j, ξ* quantifies the amplification of genetic effects across E values, *ε_j_* denotes environmental effects, and *η* quantifies the amplification of environmental effects across E values. In Scenario 1, γ_i_ will be nonzero. In Scenario 2, *ξ* will be nonzero. In Scenario 3, *ξ* and *η* will be nonzero and equal.

In this study, we apply three statistical approaches (**Figure 1b**) to UK Biobank data to detect genome-wide GxE, analyzing 33 traits (25 quantitative traits and 8 diseases; average *N*=325K) and 10 environmental variables as well as biological sex. First, we detect Imperfect genetic correlation (Scenario 1) by estimating the genetic correlation of effect sizes between sets of individuals binned on their E variables using cross-trait LD Score regression^25^ (LDSC) (**Methods**). For non-binary E variables, we estimate the genetic correlation between the most extreme quintiles of the E variable; for binary E variables, we estimate the genetic correlation between individuals in each E bin. Second, we employ PRSxE regression^28,29^, defined as a regression of the phenotype on the PRS^26,27^ multiplied by the E variable across individuals, to detect both Varying genetic variance (Scenario 2) and Proportional amplification (Scenario 3) (**Methods**); we note that PRSxE regression is not sensitive to changes in environmental variance only (**Methods**). We use PRS computed by PolyFun-pred^27^ for all analyses involving PRS. We do not standardize the E variables, and we correct for main and interaction effects of several covariates (**Methods**). Finally, we distinguish between Scenario 2 and Scenario 3 by estimating the SNP-heritability within each E bin using BOLT-REML^33^ and testing for significant differences between E bins (most extreme quintiles for non-binary E variables; each bin for binary E variables).

We assign a trait-E pair to Scenario 1 if it has a genetic correlation across E bins < 1 (regardless of whether it differs in SNP-heritability or has a significant PRSxE regression term); we assign a trait-E pair to Scenario 2 if it has both a significant PRSxE regression term and a significant difference in SNP-heritability across E bins; finally, we assign a trait-E pair to Scenario 3 if it has a significant PRSxE regression term with no significant difference in SNP-heritability across E bins (**Figure 1c**). We note that for some trait-E pairs, we detected both locus-dependent GxE (Scenario 1) and non-locus-dependent GxE (Scenario 2 or Scenario 3). In the **Supplementary Note,** we provide interpretations of test outcomes that do not correspond to exactly one of the three scenarios. We estimate the excess trait variance explained by genome-wide GxE as follows. In Scenario 1, we transform the estimate of genetic correlation across E bins to the variance scale (**Methods**; **Supplementary Note**). In Scenario 2 and Scenario 3, we approximate the relative amount of trait variance explained by GxE effects (relative to the genetic variance) as the trait variance explained by PRSxE effects divided by the trait variance explained by the PRS; this approximation is valid under a model in which the PRSxE effects are proportional to the GxE effects (**Methods**). All reported variances are transformed to the liability scale for disease traits. We have released open-source software implementing the above approaches (see Code Availability), as well as their output from this study (see Data Availability).

### Simulations

We performed simulations of the three Scenarios to evaluate the properties of the three statistical approaches. We assigned individuals to one of two E bins and simulated genetic effects at 10,000 causal SNPs based on the Scenario and E bin. We simulated sample sizes specific to each statistical approach to match our real data analyses (see below). In Scenario 1, we set the SNP-heritability to 25% and varied the genetic correlation from 99% to 94%. In Scenario 2, we set the genetic correlation to 100%, set the SNP-heritability to 25% in one E bin, and varied the SNP-heritability from 26% to 30% in the other E bin. In Scenario 3, we amplified the (genetic and environmental components of) phenotypes in one E bin by a range of values from 1.025 to 1.1. In each Scenario, we report the proportion of significant tests (P<0.05, which is fairly similar to our significance threshold for real traits; see below) for each of our three approaches: Genetic correlation (*N*=67K individuals per E bin), PRSxE regression (training *N*=337K, testing *N*=49K), and SNP-heritability by E (*N*=67K individuals per E bin). Because linkage disequilibrium (LD) does not impact GxE effects, we simulated genotypes without LD. We adjusted the methods used in our simulations accordingly. For Genetic correlation, we used cross-trait LD score regression in the special case of no LD^25^. For PRSxE regression, we used a simple shrinkage estimator in the special case of no LD to compute PRS. For SNP-heritability by E, we estimated SNP-heritability using LD score regression in the special case of no LD^36^. Further details of the simulation framework are provided in the **Methods** section.

In Scenario 1, the Genetic correlation approach reported a significant test in 93% of simulations when the true genetic correlation was 97% or smaller, whereas the PRSxE regression and SNP-heritability by E approaches were well-calibrated (**Figure 2a** and **Supplementary Table 1**). In Scenario 2, the PRSxE regression approach reported a significant test in 88% of simulations when the SNP-heritability difference was 4% or larger, and the SNP-heritability by E approach reported a significant test in more than 88% of simulations when the SNP-heritability difference was 2% or larger, whereas the Genetic correlation approach was well-calibrated (**Figure 2b** and **Supplementary Table 1**). In Scenario 3, the PRSxE regression approach reported a significant test in 88% of simulations when the proportional amplification was 1.075 or larger, whereas the Genetic correlation and SNP-heritability by E approaches were well-calibrated (**Figure 2c** and **Supplementary Table 1**). In null simulations (heritable trait with no GxE), all three statistical approaches were well-calibrated (**Supplementary Figure 1**).

**Figure 2.**
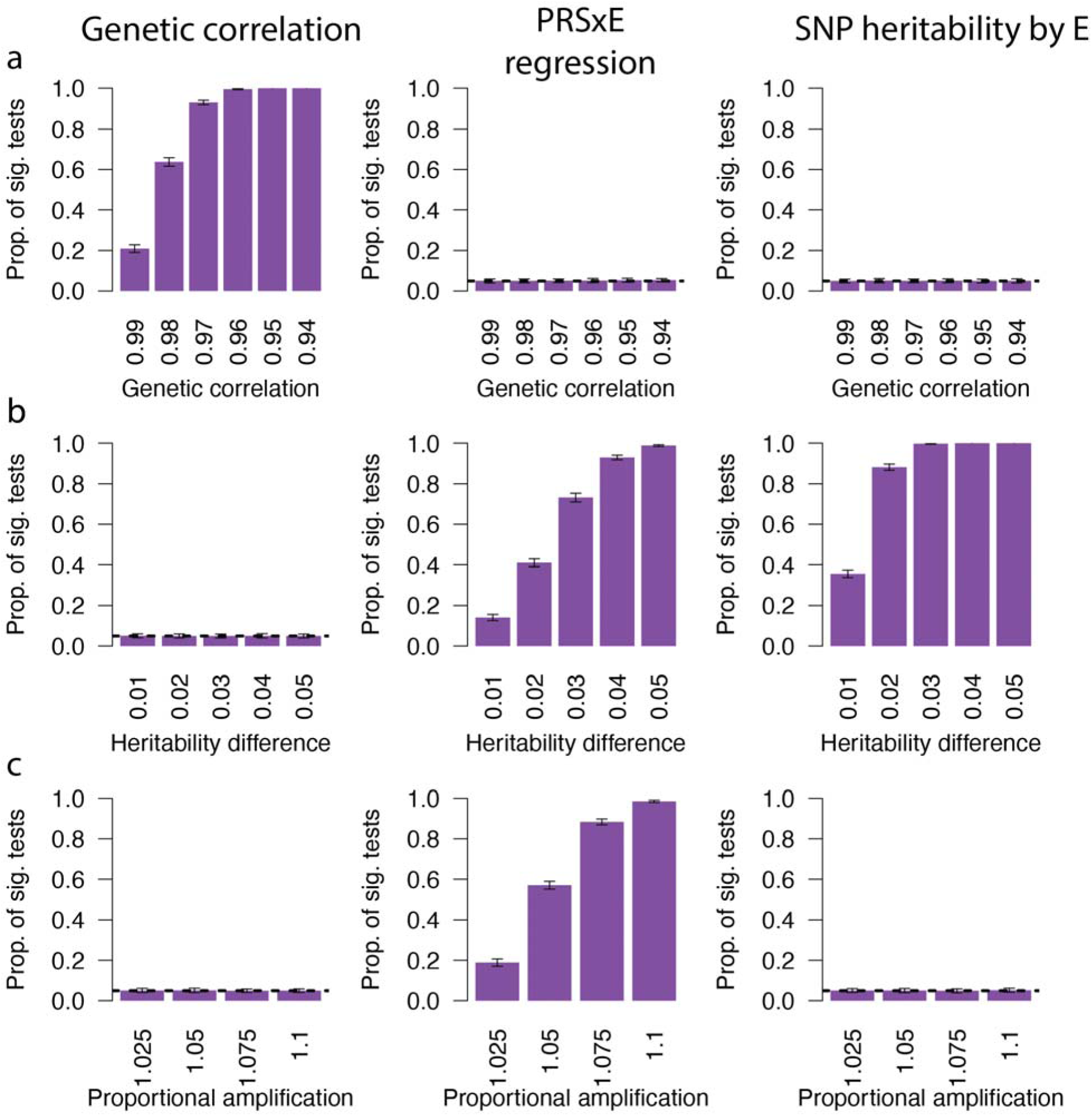
Detecting and distinguishing between 3 Scenarios of GxE interaction in simulations. Rows denote 3 Scenarios (1-3), and columns denote 3 statistical approaches. (a) Proportion of significant tests for Scenario 1 (Imperfect genetic correlation) across 3 statistical approaches. (b) Proportion of significant tests for Scenario 2 (varying genetic variance) across 3 statistical approaches. (c) Proportion of significant tests for Scenario 3 (proportional amplification) across 3 statistical approaches. Error bars denote standard deviations across 100 simulation replicates. Numerical results are reported in **Supplementary Table 1**.

We compared our framework with GxEMM^16^, a variance components-based framework that implements two GxE tests: 1) a test for polygenic GxE under homoskedasticity (GxEMM-Hom), and 2) a test for polygenic GxE under heteroskedasticity (GxEMM-Het). We note that GxEMM-Hom and GxEMM-Het do not precisely map to the 3 scenarios that we study here. In addition, because GxEMM is a variance components-based framework, it is currently unable to scale to biobank-sized datasets. We evaluated the performance of GxEMM on a sample size of 10,000 individuals, as in the simulations of ref. ^16^. We evaluated our statistical approaches using matched sample sizes, with 5,000 individuals per binary E bin and 10,000 test individuals for PRSxE regression. We kept the training data set size the same as in our main simulations (*N*=337K). In Scenario 1, the GxEMM-Hom test reported a similar proportion of significant tests as the Genetic correlation approach, whereas the GxEMM-Het test reported roughly half as many significant tests (**Supplementary Figure 2**). In Scenario 2, the GxEMM-Het test was less powerful than the PRSxE regression and SNP-heritability by E approaches, whereas the GxEMM-Hom test was well-calibrated (**Supplementary Figure 2**). In Scenario 3, the GxEMM-Het test was less powerful than the PRSxE regression approach, whereas the GxEMM-Hom test was well-calibrated (**Supplementary Figure 2**). Thus, at sample sizes that permit computational tractability, GxEMM is generally less powerful than our framework (and cannot distinguish Scenario 2 and Scenario 3).

Recognizing that “E variables” may be significantly heritable (see below), we tested whether E variables that are heritable and genetically correlated to the trait induce false positives in our PRSxE regression approach. We performed null simulations in which the E variable had SNP-heritability of 25% (matching the trait) and was 100% genetically correlated to the trait. We binned individuals based on the E variable and performed the PRSxE regression test. We determined that there was no inflation in test statistics (**Supplementary Figure 3**). Thus, the PRSxE regression is robust to E variables that are heritable and genetically correlated to the trait.

Our framework also estimates the excess trait variance explained by GxE effects, beyond what is explained by additive effects (for brevity, we refer to this as variance explained). We determined that estimates of trait variance explained were accurate in each of Scenario 1 (regression slope = 0.98; **Supplementary Figure 4a**), Scenario 2 (regression slope = 0.85; **Supplementary Figure 4b**), and Scenario 3 (regression slope = 1.05; **Supplementary Figure 4c**). We note that in both Scenario 2 and Scenario 3, G effects are correlated with GxE effects, as a correlation5etweenen genetic variance (G^2^) and the E variable implies a correlation between G and GxE. Current variance components methods do not account for this correlation and may therefore produce biased estimates of variance explained by GxE; we have verified this in simulations (**Supplementary Table 2**). Here, we report the difference in variance explained by a model including an interaction term (PRS+PRSxE terms) over a base model (PRS only) that does not include an interaction term, which is robust to this correlation (**Methods** and **Supplementary Figure 4**).

In summary, our simulations indicate that our statistical approaches attain high sensitivity and specificity in classifying trait-E pairs into the distinct scenarios of GxE considered here and produce accurate estimates of excess trait variance explained by GxE.

### Identifying gene-environment interactions across 33 complex traits/diseases and 10 E variables

We analyzed individual-level data for *N*=384K unrelated European-ancestry individuals from the UK Biobank^35^. We selected 33 highly heritable (z-score for nonzero SNP-heritability^36^ > 6) and relatively independent (squared genetic correlation^25^ < 0.5) traits and diseases (**Supplementary Table 3**). In addition, we selected 10 relatively independent E variables spanning lifestyle, diet, and other environmental exposures (*r*^2^ < 0.1; primarily from ref. ^14^; **Supplementary Figure 5**; see **Methods**). We note that these E variables are all significantly heritable, although the heritability tends to be low (mean SNP-heritability = 6%, max SNP-heritability = 15%; **Supplementary Table 4**). We assessed statistical significance using a threshold of FDR<5% across traits and E variables for a given statistical test (see **Methods**); in practice, this FDR threshold corresponded to a P-value threshold of 0.01, which is fairly similar to our simulations.

Trait-E pairs assigned to Scenario 1 (Imperfect genetic correlation) are reported in **Figure 3a** and **Supplementary Table 5**. We identified 19 trait-E pairs with genetic correlation significantly less than 1 (FDR<5%; average genetic correlation: 0.95), implicating 12 of 33 traits (including 11 quantitative traits and 1 disease) and 9 of 10 E variables tested. The implicated traits included 9 blood cell and biochemistry traits, as well as height, BMI, and asthma. On average, these interactions explained 0.30% of trait variance across all quantitative traits and 0.19% of observed-scale variance across all diseases analyzed. The lowest genetic correlation significantly less than 1 was 0.85 (se=0.06) for asthma x time spent watching television, explaining 1.5% of trait variance. The significant GxE interaction for BMI and smoking status (explaining 0.4% of trait variance) was consistent with results from ref. ^14^. Trait-E pairs assigned to Scenario 2 (Varying genetic variance) are reported in **Figure 3b** and **Supplementary Table 5**. We identified 28 trait-E pairs with significant PRSxE interaction (FDR<5%) and a significant SNP-heritability by E test (FDR<5%), implicating 13 of 33 traits (including 6 quantitative traits and 7 diseases) and 9 of 10 E variables tested. On average, these interactions explained 0.13% of trait variance across all quantitative traits and 8.8% of observed-scale variance across all diseases analyzed; the latter value is highly relevant for prediction of disease risk on the observed scale but is unlikely to reflect liability-scale GxE variance (see below and **Discussion**). Because standard interaction tests can be anti-conservative due to unmodeled heteroskedasticity^37^, we repeated our PRSxE interaction analysis using Huber-White variance estimators^38,39^ (**Methods**). We determined that results were highly concordant with our primary PRSxE interaction analysis (mean Pearson correlation in p-values for interaction across trait-E pairs: 97%; **Supplementary Table 6**), suggesting that our findings are not driven by unmodeled heteroskedasticity. Trait-E pairs assigned to Scenario 3 (Proportional amplification) are reported in **Figure 3c** and **Supplementary Table 5**. We identified 15 trait-E pairs with significant PRSxE interaction (FDR<5%) but a non-significant SNP-heritability by E test (FDR<5%), implicating 11 of 33 traits (all quantitative traits) and 9 of 10 E variables tested. On average, these interactions explained 0.17% of trait variance across all quantitative traits and 0% of observed-scale variance across all diseases analyzed.

**Figure 3.**
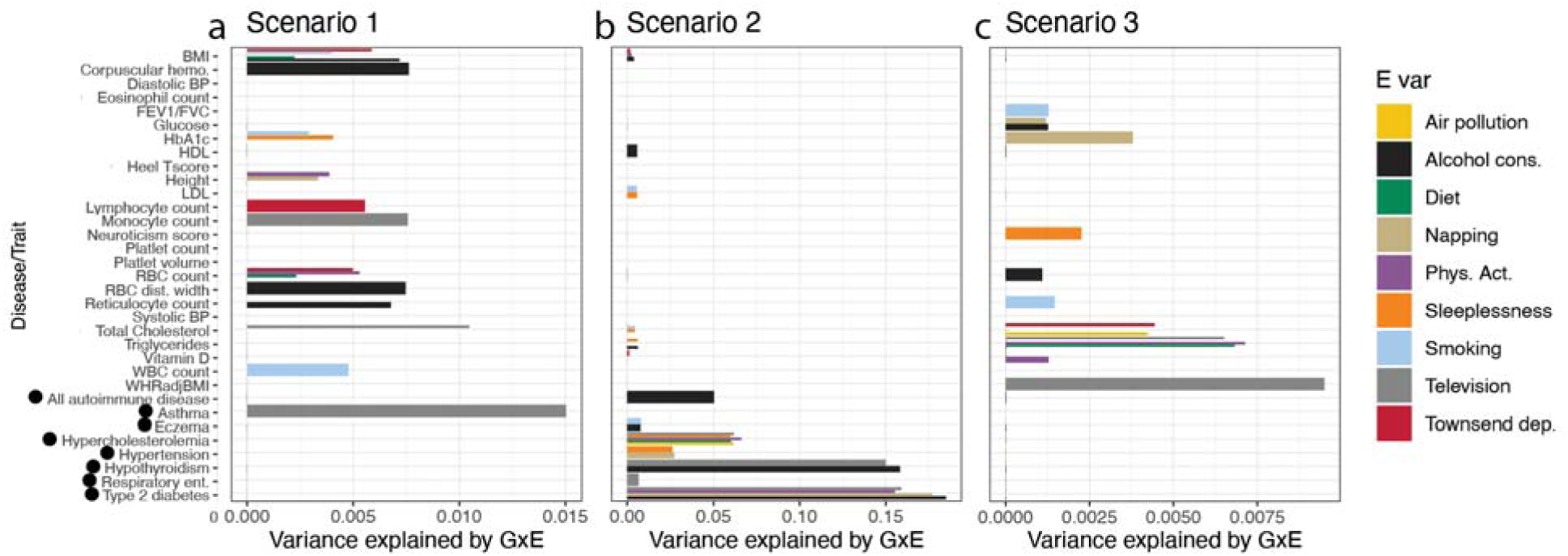
Detecting, quantifying, and distinguishing between 3 Scenarios of GxE interaction across 33 traits and 10 E variables. Traits are listed on the y-axis and estimates of excess variance explained by GxE are reported on the x-axis. Only significant results are reported (FDR < 5% across traits and E variables, computed separately for each Scenario). For traits with multiple significant E variables in a given Scenario, results for each significant E variable are reported separately using bars with smaller thickness. Disease traits are denoted with black dots. (a) Results for trait-E pairs in Scenario 1: Imperfect genetic correlation. (b) Results for trait-E pairs in Scenario 2: Varying genetic variance; we note that BMI has significant GxE for Townsend deprivation (red), physical activity (purple), and alcohol consumption (black). (c) Results for trait-E pairs in Scenario 3: Proportional amplification. Numerical results are reported in **Supplementary Table S5**.

We analyzed matched quantitative and disease traits in order to assess whether the large observed-scaled variances for diseases in Scenario 2 were recapitulated for the quantitative traits. We matched three diseases (type 2 diabetes, hypercholesterolemia, hypertension) to highly genetically correlated quantitative traits ( *r_g_* > 50 %) (HbA1c, LDL, systolic blood pressure). First, for type 2 diabetes (average observed-scale variance explained = 17% across 4 E variables; maximum of 18% for alcohol consumption), no E variable was assigned to Scenario 2 for HbA1c (**Supplementary Table 7**). Second, for hypercholesterolemia and LDL ( *r_g_* > 50 %), of the 5 E variables assigned to Scenario 2 for hypercholesterolemia (average observed-scale variance explained = 6.2% across 5 E variables; maximum of 6.6% for physical activity), only 1 E variable was assigned to Scenario 2 for LDL (average variance explained = 0.01% across 5 E variables; non-significant for physical activity) (**Supplementary Table 7**). Third, for ( *r_g_* > 0.80), of the 2 E variables assigned to Scenario 2 for hypertension (average observed-scale variance explained = 2.7% across 2 E variables; maximum of 2.7% for time spent napping), no E variable was assigned to Scenario 2 for systolic blood pressure (**Supplementary Table 7**). Thus, the large observed-scaled variances for diseases in Scenario 2 were not recapitulated for the matched quantitative traits (see **Discussion**).

We checked whether any trait-E pairs were assigned to more than one Scenario. We determined that 2 trait-E pairs were assigned to both Scenario 1 and Scenario 2 (BMI x alcohol consumption and BMI x Townsend deprivation index); 0 trait-E pairs were assigned to both Scenario 1 and Scenario 3; and 0 trait-E pairs were assigned to both Scenario 2 and Scenario 3 (which is not possible based on their definition). We also identified 108 trait-E pairs with a significant SNP-heritability by E test but non-significant PRSxE interaction (**Supplementary Table 8**); our primary interpretation is that this is due to changes in environmental variance rather than GxE interaction (**Methods**), but we cannot exclude the possibility that this is due to GxE interaction that we have incomplete power to detect.

Examples of quantitative trait-E pairs assigned to each scenario are reported in **Figure 4** and **Supplementary Table 9**. First, white blood cell count x smoking status was assigned to Scenario 1 (**Figure 4a**). The Genetic correlation approach estimated a genetic correlation between smokers and non-smokers of 0.95, which is significantly less than 1 (P=6.7e-7; FDR < 5%), explaining 0.5% of the variance of white blood cell count (vs. SNP-heritability of 30%). On the other hand, the PRSxE regression approach (P=0.46) and SNP-heritability x E approach (P=0.39) produced non-significant results. We note that smokers had 0.09 s.d. higher mean white blood cell count than non-smokers (T-test P<1e-16), as previously reported^40^. Second, BMI x physical activity was assigned to Scenario 2 (**Figure 4b**). The PRSxE regression approach (P=4.6e-5) and SNP-heritability x E approach (SNP-heritability of 0.38 for highest E quintile vs. 0.33 for lowest E quintile; P<7e-4) both produced significant results (FDR < 5%), explaining 0.16% of the variance of BMI (vs. SNP-heritability of 33%). On the other hand, the genetic correlation approach produced a non-significant result (P=0.43). We note that BMI and physical activity were correlated (r=-0.09, P<1e-16) as previously reported^41^. Third, WHRadjBMI x time spent watching television (TV time) was assigned to Scenario 3 (**Figure 4c**). The PRSxE regression approach produced a significant result (P=5e-3; FDR < 5%), explaining 0.95% of the variance of WHRadjBMI. On the other hand, the genetic correlation approach (P=0.29) and SNP-heritability x E approach (P=0.08) produced non-significant results. We note that WHRadjBMI and TV time were correlated (*r* = 0.08, P<1e-16).

**Figure 4.**
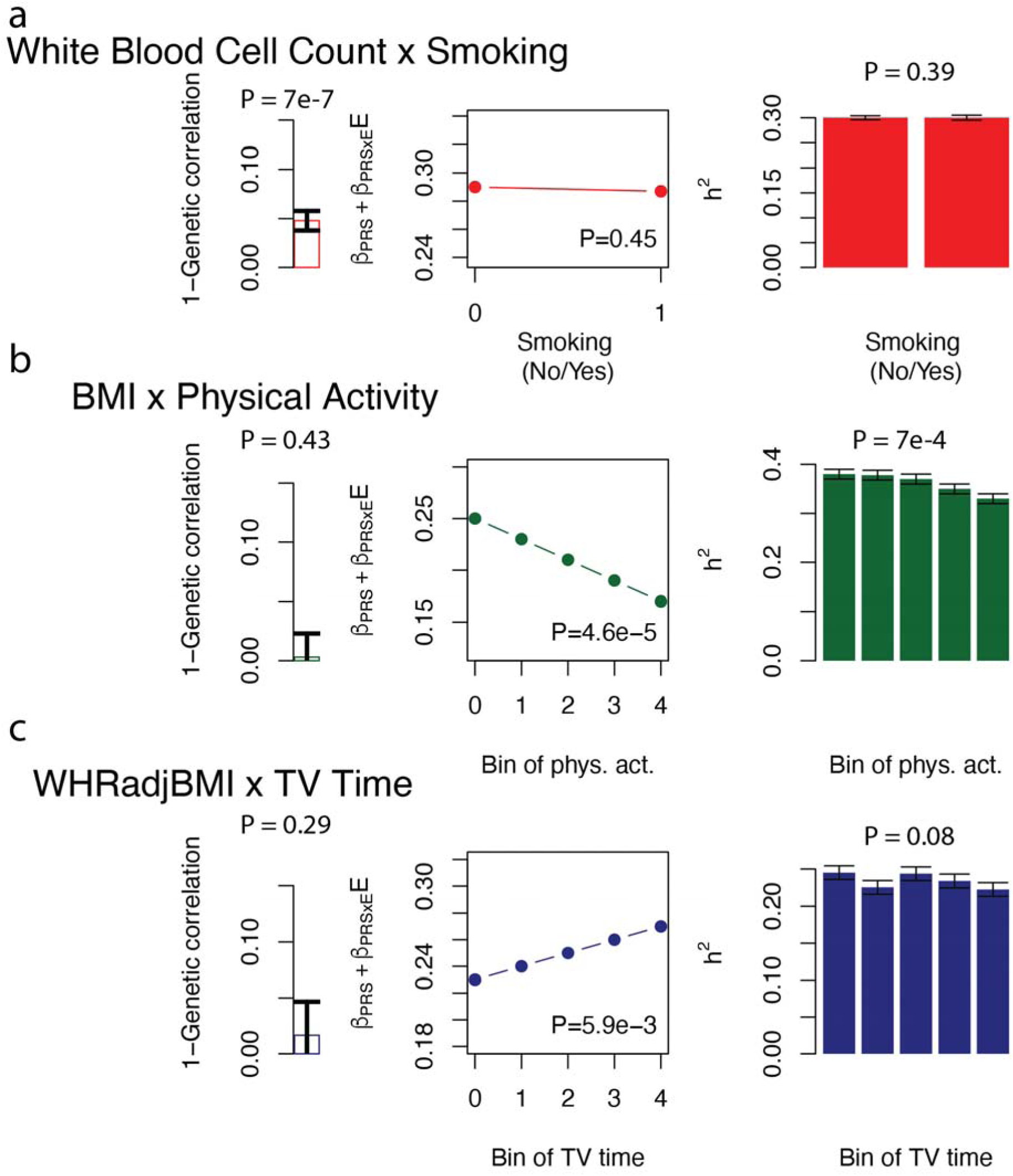
Examples of 3 Scenarios of GxE interaction. (a) White blood cell count x smoking status is consistent with Scenario 1: Imperfect genetic correlation. (b) BMI x physical activity is consistent with Scenario 2: Varying genetic variance. (c) Waist-to-hip ratio adjusted for BMI x Time spent watching TV is consistent with Scenario 3: Proportional amplification. Numerical results are reported in **Supplementary Table S8**.

In summary, we detected GxE interaction in each of the three scenarios across the 33 traits and 10 E variables analyzed. We estimate that these GxE effects explain 0.6% of trait variance across all quantitative traits and 9.0% of observed-scale variance across all diseases analyzed, compared to average SNP-heritability of 29% (s.e. 3% across traits).

### Identifying gene-sex interactions across 33 diseases/complex traits

We analyzed the same 33 traits for GxSex interaction using the same 3 statistical approaches. Traits assigned to Scenario 1 (Imperfect genetic correlation) are reported in **Figure 5a** and **Supplementary Table 10**. We identified 18 quantitative traits and 4 diseases with cross-sex genetic correlation significantly less than 1 (FDR<5%; average genetic correlation: 0.92), consistent with previous results^22^. On average, these interactions explained 2.6% of trait variance across all quantitative traits and 2.4% of observed-scale variance across all diseases analyzed. The lowest significant genetic correlation was 0.66 for WHRadjBMI^22^, explaining 17% of trait variance. Traits assigned to Scenario 2 (Varying genetic variance) are reported in **Figure 5b** and **Supplementary Table 10**. We identified 6 quantitative traits and 6 diseases with significant PRSxSex interaction (FDR<5%) and a significant SNP-heritability by Sex test (FDR<5%). On average, these interactions explained 0.13% of trait variance across all quantitative traits and 5.6% of observed-scale variance across all diseases analyzed; the latter value is highly relevant for prediction of disease risk on the observed scale but is unlikely to reflect liability-scale GxSex variance (see below and **Discussion**). Traits assigned to Scenario 3 (Proportional amplification) are reported in **Figure 5c** and **Supplementary Table 10**. We identified 8 quantitative traits and 0 diseases with significant PRSxSex interaction (FDR<5%) but a non-significant SNP-heritability by Sex test (FDR<5%). On average, these interactions explained 0.06% of trait variance across all quantitative traits (a very small contribution) and 0% of observed-scale variance across diseases analyzed. Of the 30 traits implicated across three scenarios, we identified 7 traits assigned to both Scenario 1 and Scenario 2, and 5 traits assigned to both Scenario 1 and Scenario 3 (**Supplementary Table 10**). We also identified 2 traits with a significant SNP-heritability x Sex test but non-significant PRSxSex interaction (**Supplementary Table 11**); our primary interpretation is that this is due to changes in environmental variance rather than GxSex interaction (**Methods**).

**Figure 5.**
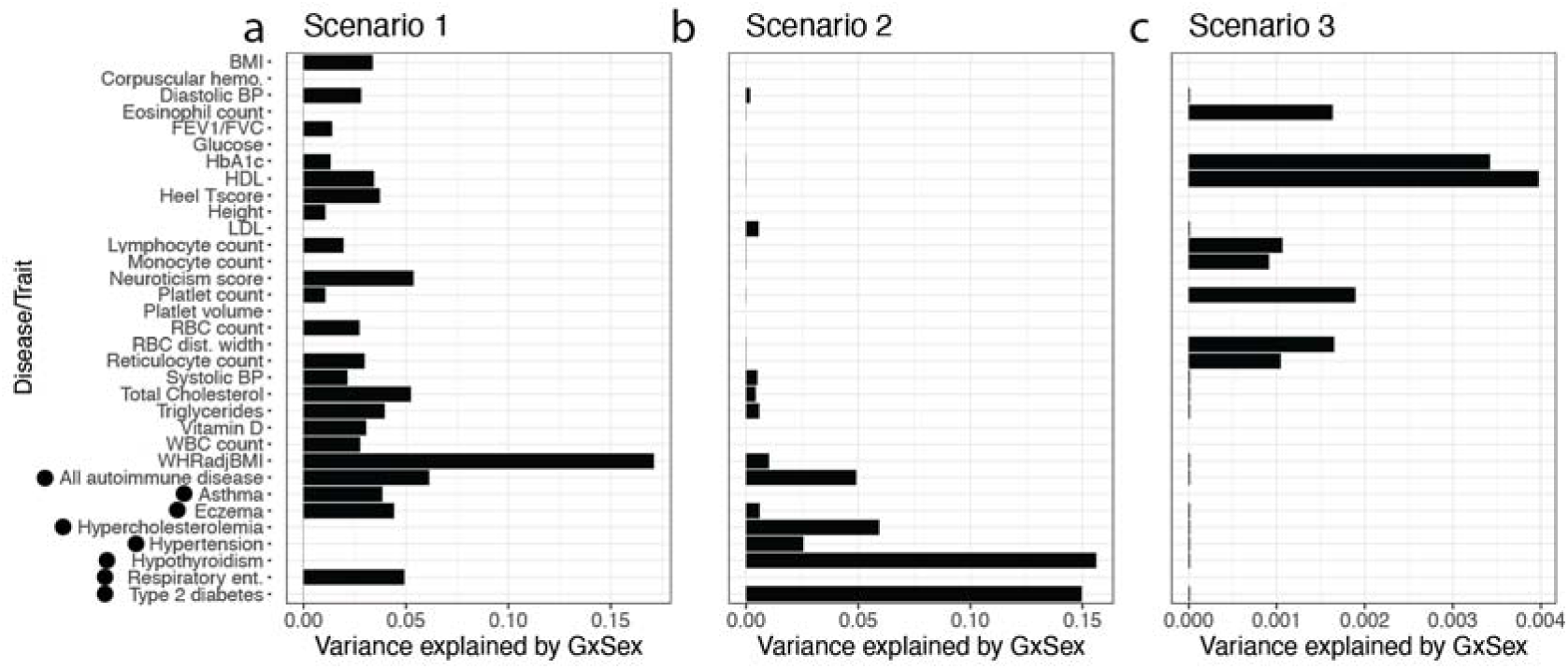
Detecting, quantifying, and distinguishing between 3 Scenarios of GxSex interaction across 33 traits. Traits are listed on the y-axis and estimates of excess variance explained by GxSex are reported on the x-axis. Only significant results are reported (FDR < 5% across traits, computed separately for each Scenario). Disease traits are denoted with black dots. (a) Results for traits in Scenario 1: Imperfect genetic correlation. (b) Results for traits in Scenario 2: Varying genetic variance. (c) Results for traits in Scenario 3: Proportional amplification. Numerical results are reported in **Supplementary Table S9**.

We again analyzed matched quantitative and disease traits to assess whether the large observed-scaled variances for diseases in Scenario 2 were recapitulated for the quantitative traits. First, for type 2 diabetes and HbA1c ( *r_g =_* _0.66_), type 2 diabetes was assigned to Scenario 2 (observed-scale variance explained = 15%), but HbA1c was not assigned to Scenario 2 (**Supplementary Table 12**). Second, for hypercholesterolemia and LDL ( *r_g =_* _0.71_) hypercholesterolemia was assigned to Scenario 2 (observed-scale variance explained = 5.9%), and LDL was also assigned to Scenario 2 but with much lower variance explained (0.56%) (**Supplementary Table 12**). Third, for hypertension and systolic blood pressure (*r_g =_* _0.80)_, hypertension was assigned to Scenario 2 (observed-scale variance explained = 2.5%), and systolic blood pressure was also assigned to Scenario 2 but with much lower variance explained (0.50%) (**Supplementary Table 12**). Thus, the large observed-scaled variances for diseases in Scenario 2 were not recapitulated for the matched quantitative traits (see **Discussion**).

Examples of quantitative traits with significant GxSex assigned to each Scenario are reported in **Figure 6** and **Supplementary Table 13**. First, neuroticism score was assigned to Scenario 1 (**Figure 6a**). The Genetic correlation approach estimated a cross-sex genetic correlation of 0.90, which is significantly less than 1 (P=3.5e-9; FDR < 5%), explaining 5.0% of the variance of neuroticism score. On the other hand, the PRSxSex regression approach (P=0.58) and SNP-heritability by Sex approach (P=0.45) produced non-significant results. We note that males had lower prevalence of neuroticism than females (1.6% vs. 2.3% in top score for neuroticism, P<1e-16), as previously reported^42^. Second, systolic blood pressure was assigned to Scenario 2 (**Figure 6b**). The PRSxSex regression approach (P=3.8e-5) and SNP-heritability by Sex approach (SNP-heritability of 32% for males and 27% for females; P=2e-15) both produced significant results (FDR < 5%), explaining 0.14% of the variance of systolic blood pressure (vs. SNP-heritability of 28%). In addition, the genetic correlation approach produced a significant result (P=7e-5), explaining 2.5% of the variance of systolic blood pressure (Scenario 1); this implies that multiple types of GxSex interaction impact systolic blood pressure. We note that males had higher systolic blood pressure than females (P<2e-16), as previously reported^43^. Third, HDL cholesterol was assigned to Scenario 3 (**Figure 6c**). The PRSxSex regression approach produced a significant result (P<2e-16; FDR < 5%), explaining 0.4% of the variance of HDL cholesterol. On the other hand, the SNP-heritability by Sex approach (P=0.09) was not significant. However, the genetic correlation approach estimated a cross-sex genetic correlation of 0.93, which is significantly less than 1 (P=5e-6; FDR < 5%), explaining 3.5% of the variance of HDL cholesterol (Scenario 1); this implies that multiple types of GxSex interaction impact HDL cholesterol. We note that males had 0.83 s.d. lower HDL cholesterol than females (P<1e-16).

**Figure 6.**
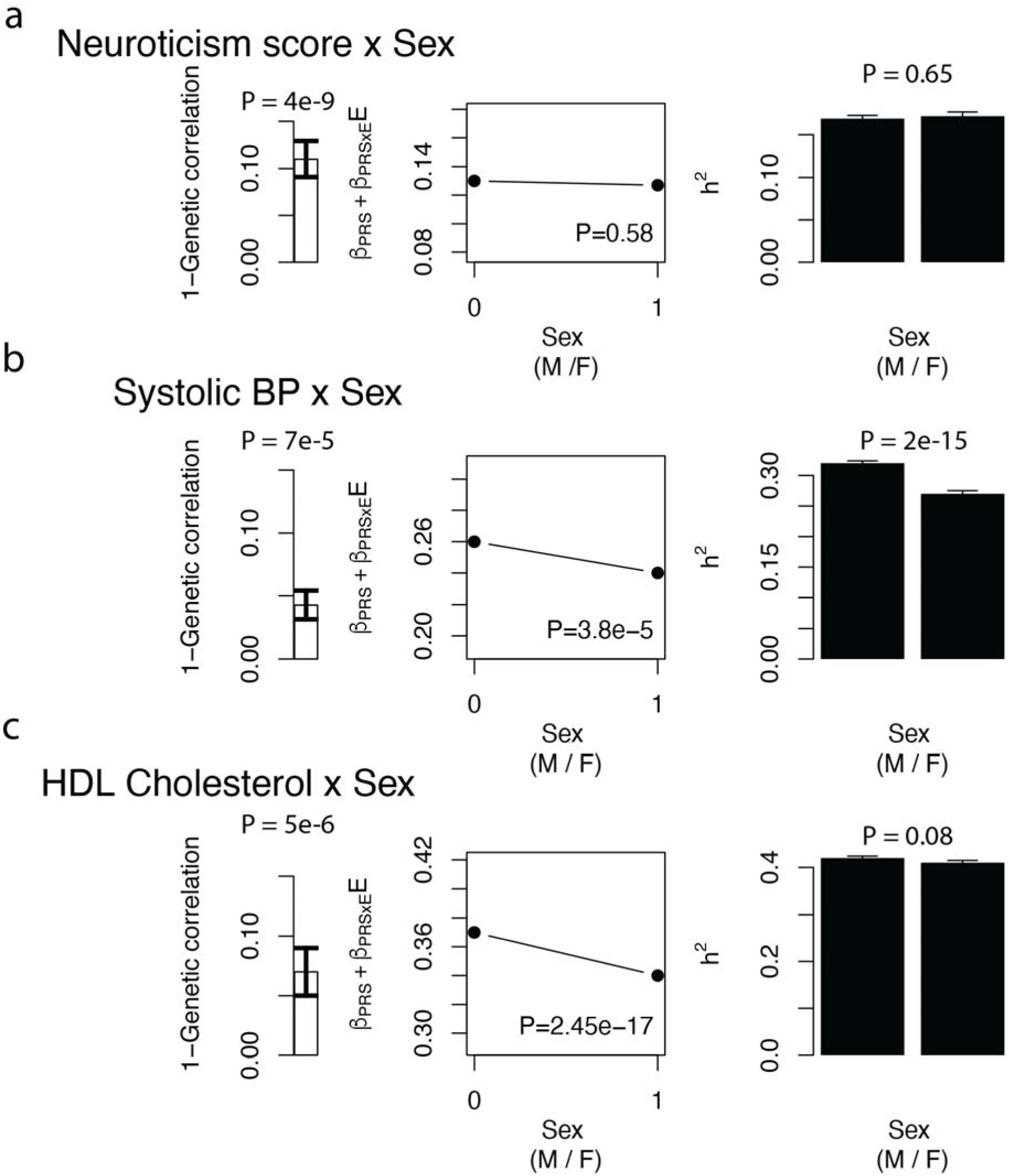
Examples of 3 Scenarios of GxSex interaction. (a) Neuroticism score x Sex is consistent with Scenario 1: Imperfect genetic correlation. (b) Systolic blood pressure x Sex is consistent with Scenario 1 and Scenario 2. (c) HDL Cholesterol x Sex is consistent with Scenario 1 and Scenario 3. Numerical results are reported in **Supplementary Table S11**.

In summary, we detected GxSex interaction in each of the three scenarios across the 33 traits analyzed. We estimate that these GxSex effects explain 2.8% of trait variance across all quantitative traits and 8.0% of observed-scale variance across all diseases analyzed, compared to average SNP-heritability of 29% (s.e. 3%).

## Discussion

We have applied three statistical approaches to detect, quantify, and distinguish the genome-wide contributions of three different types of GxE interaction (**Figure 1a**) across 33 UK Biobank traits, analyzing 10 E variables spanning lifestyle, diet, and other environmental exposures as well as biological sex. We determined that GxE interactions (involving these E variables) and GxSex interactions each explained a significant fraction of phenotypic variance, representing an appreciable contribution to trait architectures. It is possible that GxE interactions involving E variables not studied here could explain even more phenotypic variance. However, the much higher (observed-scale) variance explained by GxE and GxSex effects (in Scenario 2) for disease traits than for quantitative traits is complicated by scale effects (see below).

Our finding of distinct explanations underlying GxE interactions (**Figure 1a**) motivates a unified model consistent with this finding. We propose a model in which GxE occurs at different levels of a hierarchy that leads from genetic variants to pathways to complex traits or disease (**Supplementary Figure 6**). In this model, Scenario 1 (Imperfect genetic correlation) occurs when an E variable modifies the effects of individual variants (or sets of variants), differentially impacting different parts of the genome; Scenario 2 (Varying genetic variance) occurs when an E variable modifies all of the pathways underlying genetic risk, uniformly impacting genetic variance; and Scenario 3 (Proportional amplification) occurs when an E variable modifies all aspects of trait biology, proportionately impacting both genetic and environmental variance. Under this model, an E variable can modify any point along the hierarchy from genetic variants to pathways to complex traits or disease. Further investigation and validation of this model is a direction for future research.

Our study represents an advance over previous studies investigating genome-wide GxE. First, we distinguish three different types of GxE interaction: Imperfect genetic correlation, Varying genetic variance, and Proportional amplification (**Figure 1a**; also see **Supplementary Figure 6**). Second, most variance components methods for detecting genome-wide GxE^13–15,17,18^ cannot detect genome-wide GxE unless SNP-heritability varies across E bins (Scenario 2). An exception is GxEMM^16^, which detects other types of GxE by explicitly modeling genetic and environmental variance that varies with the E variable; however, GxEMM is less computationally tractable and generally less powerful than our framework (**Supplementary Figure 2**). Third, variance components methods that assume independence between G and GxE effects are susceptible to bias if G and GxE effects are correlated, but our statistical approaches are robust to this possibility (**Supplementary Figure 4**. Fourth, previous methods have not been applied at biobank scale across a broad range of traits; the statistical approaches that we propose are computationally scalable to very large data sets (see **Methods**), enabling our biobank-scale analyses implicating 60 trait-E pairs with significant GxE and 30 traits with significant GxSex. Fifth, a recent study reported that GxSex acts primarily through amplification^24^ (Scenario 2 and Scenario 3), but our analyses of GxSex determined that Imperfect genetic correlation (Scenario 1) explained a larger proportion of trait variance than amplification; in addition, ref. ^24^ did not estimate contributions to trait variance and did not distinguish between Scenario 2 and Scenario 3, as we do here.

Our study has several implications. First, our results narrow the search space of traits and E variables for which genome-wide association studies of GxE interactions are most likely to be fruitful; in particular, trait-E pairs with substantial trait variance explained by Scenario 1 (Imperfect genetic correlation) (**Supplementary Table 5**) should be prioritized for locus-specific analyses, in preference to trait-E pairs with trait variance explained by Scenario 2 or Scenario 3. Second, our results imply that there is broad potential to improve polygenic risk scores (PRS) by leveraging information on E variables in training and/or test samples^44^. Third, there is broad potential to prioritize individuals for which a lifestyle intervention to modify an E variable would be most effective based on their genetic profile. Fourth, previous work has suggested that population-specific causal effect sizes in functionally important regions may be caused by GxE^45^, motivating efforts to partition the imperfect genetic correlations across E bins that we have identified across functionally important regions. Fifth, the significant contribution of GxE to trait architectures—even when restricting to the limited set of E variables that we analyzed here—implicates GxE effects as a factor in “missing heritability”, defined as the gap between estimates of SNP-heritability^30^ and estimates of narrow-sense heritability^46^ (e.g. from twin studies^47^); although GxE effects are not included in the *definition* of narrow-sense heritability, they can inflate twin-based *estimates* of narrow-sense heritability, analogous to GxG effects^48^. All of these implications motivate directions for future research.

Our study has several limitations. First, our analyses assess GxE and GxSex interaction for disease traits on the observed scale (and then transform estimates to the liability scale), consistent with prevailing approaches for variance component analysis of disease traits^31–34^. Our analyses of matched quantitative and disease traits (**Supplementary Table 7**, **Supplementary Table 12**) strongly suggest that the much higher (observed-scale) variance explained by GxE and GxSex effects (in Scenario 2) for disease traits than for quantitative traits (**Supplementary Table 14;** also see **Figure 3b** and **Figure 5b**) is a consequence of analyzing disease traits on the observed scale, and unlikely to reflect liability-scale GxE or GxSex variance; directly modeling GxE or GxSex interaction on the liability scale^50,51^ is an important direction for future research. Nonetheless, these observed-scale interactions are likely to be highly relevant for prediction of disease risk on the observed scale. Second, the E variables that we analyzed comprise an extremely limited subset of the set of E variables that may contribute to GxE effects (and their values may be subject to measurement error); even when GxE effects are detected, the implicated E variable may be tagging an unmeasured causal E variable with larger GxE effects. Third, our use of PRSxE regression to detect GxE is limited by the accuracy of PRS and may require larger training sample sizes (enabling more accurate PRS) to be well-powered, particularly for less heritable traits. The average accuracy of the PRS across traits in the held-out set of 49K individuals was 9.2%, as measured by *r*^2^ between predicted and true phenotypes (**Supplementary Table 3**). Fourth, our estimates of the trait variance explained by GxE effects detected via PRSxE analyses assume that PRSxE effects extrapolate linearly to GxE effects (**Methods**); we believe that this is a reasonable assumption, but we cannot formally exclude the possibility that genetic effects captured by PRS interact differently with an E variable than genetic effects not captured by PRS. Fifth, most of the E variables that we study are weakly heritable (**Supplementary Table 4**), raising the possibility of GxG (rather than GxE) effects; we consider GxG to be an unlikely explanation given the E variables’ low SNP-heritabilities, but we cannot formally exclude this possibility. Sixth, our use of PRSxE regression to detect GxE may be anti-conservative due to unmodeled heteroskedasticity^37^; however, we obtained nearly identical results using Huber-White variance estimators (also known as robust regression^38,39^) (**Supplementary Table 6**), suggesting that this does not impact our findings. We note that we observe many instances of differences in trait variance across E variables (**Supplementary Table 8**), but these alone are not indicative of GxE interactions. Seventh, our use of PRSxE regression to detect GxE may produce false positives if there is a nonlinear relationship between E and trait value; we included an E^2^ term in PRSxE regressions to ameliorate this possibility but determined that inclusion or exclusion of the E^2^ term had little impact on our results (**Supplementary Table 15**), suggesting that nonlinear effects do not greatly impact our findings. Eighth, we have analyzed British-ancestry samples from the UK Biobank, but an important future direction is to extend our analyses to cohorts of diverse genetic ancestry^52,53^, which may differ in their distributions of E variables, tagging of causal E variables by measured E variables, and/or causal GxE effects (analogous to differences in main G effects^45,54).^ Eighth, we do not analyze GxAge interaction (and we note the limited age variation in UK Biobank samples; age = 55 ± 8 years), but we highlight GxAge interaction and longitudinal data as important directions for future research^51,55,56^. Despite these limitations, our work quantifies and distinguishes three different types of GxE interaction across a broad set of traits and E variables.

## Code Availability

Cross trait LDSC: https://github.com/bulik/ldsc

BOLT-LMM: https://alkesgroup.broadinstitute.org/BOLT-LMM/downloads/

PRSxE regression: Will be added upon publication.

Code to reproduce analysis: Will be added upon publication.

## Data Availability

We will make the results of the three statistical approaches we use here publicly available upon publication.

## Data Availability

All data produced in the present work are contained in the manuscript.

## Acknowledgements

We are grateful to Martin Zhang, Ben Strober, Xilin Jiang, and Jordan Rossen for helpful discussions and Sriram Sankararaman and Ali Pazokitoroudi for comments on an earlier version of this manuscript. This research was conducted using the UK Biobank resource under application no. 16549 and funded by National Institutes of Health (NIH) grants R01 MH101244, R37 MH107649 and R01 HG006399. The funders had no role in study design, data collection and analysis, decision to publish or preparation of the manuscript.

## Methods

### Data sources and preprocessing

We used data from the UK Biobank in all our analyses. For polygenic score-based analyses that required a training and testing dataset, we used a set of 337K unrelated white British individuals for training^27^. For testing, we used a set of 49K European individuals who are unrelated to each other and to the training cohort^27^. Note that while “testing” typically refers to a setting where the ultimate goal is to assess PRS accuracy, here we use it to refer to the set of samples we in which we run a regression of phenotype on PRSxE and covariates. We used polygenic scores generated by ref. ^27^. We used the linear scoring function in Plink v1.9^57^ to compute polygenic scores in the set of 49K test individuals.

### Choice of traits and environmental variables

We chose a set of 33 traits with SNP heritability Z scores > 6 and squared genetic correlation less than 0.5 (**Supplementary Table 3, Supplementary Table 4**). We chose a set of 10 E variables, including 5 previously analyzed E variables from ref. ^14^ and 5 additional E variables (Air pollution, time spent napping, sleeplessness, Diet, wheat consumption) (**Supplementary Figure 5)**.

To compute the Diet variable, we performed PCA on a covariance matrix consisting of several diet variables: cooked vegetable intake, salad intake, fresh fruit intake, processed meat intake, poultry intake, beef intake, pork intake, coffee intake (**Supplementary Figure 7**). We used the function prcomp in R and extracted the first PC.

### Genetic correlation approach to detecting GxE

We performed GWAS using BOLT-LMM^26^ within bins of E variables. Then, we used bivariate LD Score regression^36^ to estimate the genetic correlation between the top and bottom quintiles of E variables; for binary E variables, we estimated the genetic correlation between individuals in each E bin. We used imputed SNPs with MAF > 0.01% and used the --no-intercept option to increase our power. Computed a Z score testing against the null hypothesis that the genetic correlation is 1 as:

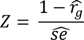

### PRSxE regression approach to detecting GxE

Our PRSxE regression takes the following form:

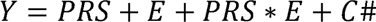

where Y is the trait value, PRS is the polygenic score for the trait (see *Data sources and preprocessing*), E is the environment variable, and C is a set of covariates. For all analyses, we correct for the following covariates: age, sex, 10 genetic PCs computed in the held-out set, the squared E variable: E^2^, age*sex, E*age, E*sex. We carried out this regression using the Python package statsmodels v0.14. We also compute a ‘base’ model, which is the same regression without the PRSxE term. We use the p-value associated with the PRSxE term in the interaction model to assess significance.

To test whether our results were driven by heteroskedasticity, we performed the same analysis using robust standard errors as implemented in statsmodels using the ‘H1’ covariance matrix (**Supplementary Table 12**).

We note the PRSxE regression test is not expected to produce a significant finding if the environmental variance changes as a function of E but the genetic variance does not change as a function of E, because the PRS does not measure changes in environmental variance.

### SNP-heritability by E approach to detecting GxE

We used BOLT-REML^33^ v2.3.6 to compute heritability in bins of E variables. To test for a significant difference in heritability between two bins, we computed a Z score as:

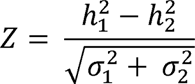

where 1 and 2 index the E bins.

### False Discovery Rate (FDR) control

We chose a 5% FDR control separately for each statistical approach (Genetic correlation, PRSxE regression, and SNP-heritability by E) using the qvalue R package^58^. We ensured our one-sided test against a null genetic correlation of 1 did not produce a skewed P-value distribution, which could indicate improper choice of a one-sided test. We chose to control the FDR separately for GxE and GxSex analyses because we expected the proportional of truly null tests to be different between GxE and GxSex. In particular, we expected to find more truly positive GxSex tests given previous studies^22,24^. Consistent with this, we found the qvalue procedure for estimating the proportion of truly null hypotheses failed in the GxSex analyses and we had to set the proportion of true null tests (π_0_) to 1, which is equivalent to the Benjamini-Hochberg procedure^59^. Story and Tibshirani^60^ argue this is much more conservative than the qvalue procedure. Our choice to control each E variable together is conservative, but accounts for non-zero correlations between E variables.

### Classification of trait-E pairs into Scenarios

We combined the results of the three statistical approaches to classify trait-E pairs into 3 distinct scenarios. We classified trait-E pairs into Scenario 1 if the Genetic correlation was significantly less than 1. We classified trait-E pairs into Scenario 2 if the SNP-heritability by E and PRSxE regression approaches were significant. We classified trait-E pairs into Scenario 3 if the PRSxE regression approach was significant but the SNP-heritability by E approach was not significant. It is possible that the SNP-heritability by E approach is significant but the PRSxE regression approach is not significant, which should not be viewed as an instance of GxE because the SNP-heritability difference may be driven by changes to the environmental variance rather than the genetic variance. In addition, it is possible for trait-E pairs to be classified into both Scenario 1 and Scenario 2, or both Scenario 1 and Scenario 3, but not both Scenario 2 and Scenario 3 because significance or non-significance of the SNP-heritability by E approach are mutually exclusive.

### Scalability of statistical approaches

We consider the scalability of the three statistical approaches we use here. First, there is the computational cost of producing the input to our statistical approaches. For the genetic correlation test, this consists of running GWAS in bins of E variables. There are many scalable approaches for this, including BOLT-LMM^26^, regenie^61^, and fastGWA^62^. For PRSxE regression, this consists of computing PRS weights. There are many scalable approaches for this including BOLT-LMM^26^, PRScs^63^, SBayesR^64^, and LDpred2^65^. SNP-heritability by E does not require generating additional input. In these analyses, we use BOLT-LMM for GWAS, which has a runtime that scales with O(MN), where M is the number of SNPs and N is the sample size of individuals. For PRSxE regression we use weights computed by Weissbrod et al 2022^27^, who did not publish an analysis of runtime. Second, there is the computational cost of the statistical approaches themselves. For genetic correlation, we use cross-trait LDSC^25^, which runs in seconds (< 30s for the SNP sets that we analyze here). For PRSxE regression, we use a multiple regression, which also runs in seconds (< 30s for the sample size that we analyze here). For SNP-heritability by E, we use BOLT-REML^33^, which has a runtime that scales with O(MN).

### Simulations

To test the power of each approach, we simulated 1,000 replicates of each scenario. In all cases, we simulated two E bins and varied the parameters according to the respective generative models. For each replicate, we simulated *M*=10,000 causal SNPs with effect sizes drawn from a specified distribution. We generated unlinked genotypes with binomial sampling from an allele frequency of 0.5.

We simulated causal effect sizes for each scenario as follows:

Scenario 1

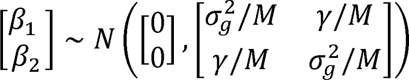

We simulated 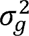 = 0.25 and varied γ to produce genetic correlations *γ_g_* ∈{1,0.99,0.98,0.97,0.96,0.95,0.94}.

Scenario 2

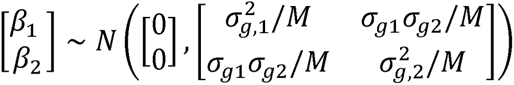

We simulated 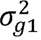 = 0.25 and set 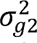 to produce a a difference in heritability: {0,0.01,0.02,0.03,0.04,0.05}.

Our choice of covariance ensures the genetic correlation is one.

Scenario 3

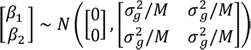

We set 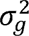 = 0.25 To simulate proportional amplification, we multiplied the phenotypes for individuals in environment 2 by a constant: ={1.0,1.025,1.05,1.075,1.1}.

Using the simulated causal effect sizes, we simulated GWAS effect size estimates as:

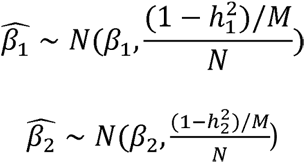

where 1 and 2 index the environments and *N* denotes GWAS sample size. We estimate 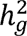 from the simulated causal effect sizes by first computing the χ2 statistic (χ2:= *N \** 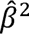), then computing, 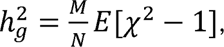 where *E* denotes the mean computed over the independent SNPs^25^.

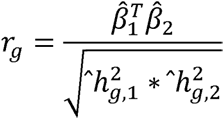

where *T* denotes the transpose. We compute standard errors for the estimates using a jackknife over SNPs, where we leave out one SNP at a time because they are independent.

To simulate PRSxE, we first simulated causal effect sizes for 10,000 independent SNPs. Then, we compute PRS weights analytically as:

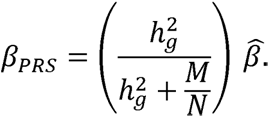

This simple shrinkage estimator can be interpreted as the posterior mean causal effect size under a normal prior (in the special case of no LD), and is similar to the posterior mean causal effect size under a point-normal prior (in the special case of no LD) when the genetic architecture is highly polygenic^66^, as simulated here.

We estimate 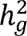 without knowledge of the E bins, mimicking estimation of SNP-heritability across the 337K individuals; we estimate 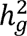 as the sum of squared standardized effect sizes (averaged across E bins).

We also evaluated the performance of GxEMM in detecting GxE in Scenarios 1, 2, and 3. We followed the simulation framework in the original publication and simulated 1,000 causal SNPs and 10,000 individuals. We simulated a binary E variable and drew SNP effects according to each Scenario. We performed two tests within the GxEMM framework: 1) IID versus Hom, which tests for polygenic GxE under homoscedasticity, and 2) Free versus Hom, which test for polygenic GxE allowing for heteroskedasticity. We performed a Wald test as implemented in GxEMM and compared the point estimates of heritability in the free model to the simulated heritability in each of the environments. We performed 100 simulation replicates.

To compare GxEMM with our tests, we simulated data under the same framework with matched sample sizes. Specifically, for genetic correlation and SNP-heritability x E, we simulated 5,000 individual per E bin (total N=10,000). For PRSxE, we used a training set sample size of 337K, which matches the real data, and a held out set of N=10,000.

### Estimation of trait variance explained

For trait-E pairs in Scenario 1, we compute the trait variance explained by GxE as (1 - *r_g_*/2) for binary E variables (where *r_g_* is the genetic correlation between the two E bins) (**Supplementary Note**) and 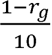 for continuous E variables (where *r_g_* is the genetic correlation between the top and bottom quintiles of E values). To obtain the transformation for continuous E variables, we used our simulations (see above) to examine the relationship between estimated genetic correlation and the variance explained by GxE. We found when we binned the E variable into 5 bins and computed the genetic correlation between the top and bottom bins, the transformation 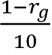 produced accurate estimates of the variance explained by GxE. For trait-E pairs in Scenarios 2 or 3, we divide the variance explained by the PRSxE regression term by the variance explained by the PRS and multiply by the SNP-heritability. We verified these scaling procedures produce accurate estimates of the excess variance explained by GxE in simulations (**Supplementary Figure S4**). For Scenario 1, we simulated a continuous E variable with mean 0 and variance 1 for 337K individuals. We simulated main genetic effects drawn from a normal distribution with mean 0 and variance 0.25 and environment interaction effects from a normal distribution with mean 0 and variance across a range of parameters (1e-1 to 1e-5) for 5,000 SNPs. We binned individuals into 5 bins and ran a GWAS in the top and bottom bins and compute the genetic correlation between the bins. Then, we scaled the estimates according to the formula above (**Supplementary Figure S4a)**. For Scenarios 2 and 3, we simulated 1,000 causal SNPs from a normal distribution with mean 0 and variance 0.25. We simulated a continuous E variable with mean 0 and variance 1 for 49K individuals. We set the amplification parameter to 0.1 and generated phenotypes according to Eq. 1 (*Overview of methods*). We performed GWAS and estimated PRS weights as in the *Simulations* section. Then, we ran the PRSxE test and computed the variance explained. We then compared this to the true variance explained (**Supplementary Figure S4b, S4c**). This scaling assumes that PRSxE effects linearly extrapolate to GxE effects. We do not use the estimates of differences in SNP-heritability by E to estimate the variance explained by GxE. When reporting average variance explained per trait, we computed the R^2^ for each trait using a model including all marginally significant (FDR < 5%) interaction terms for that trait (**Supplementary Table 14**).

## Supplementary Material

**Figure S1.**
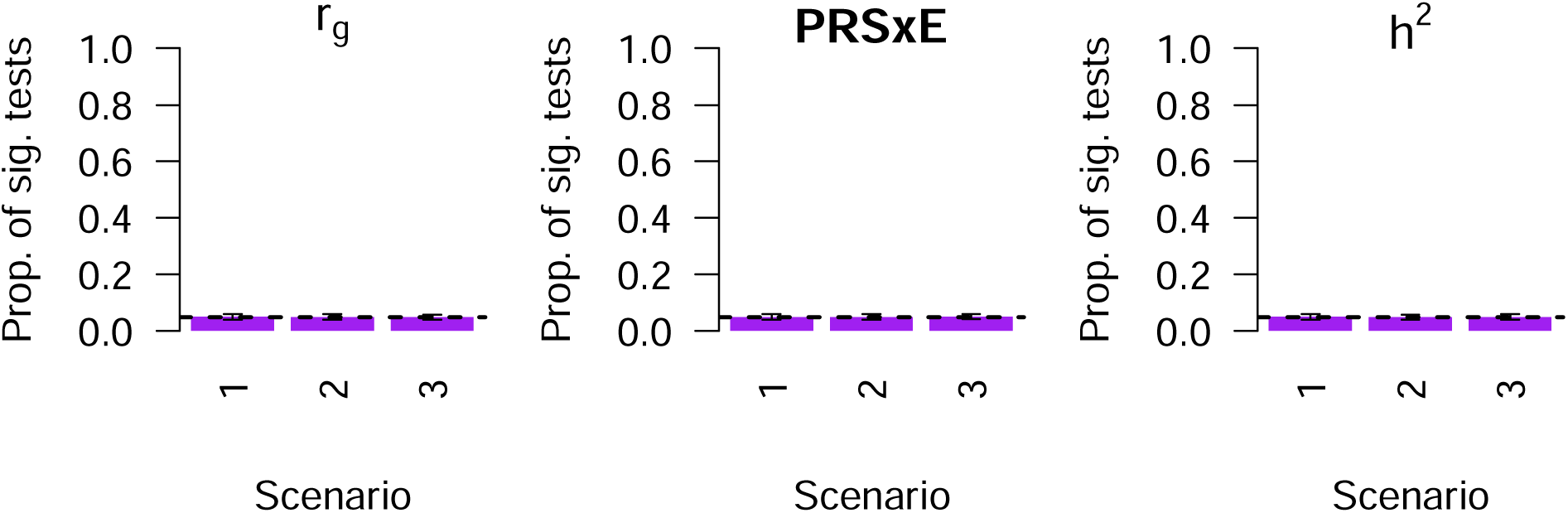
Results of 3 statistical approaches for detecting GxE in null simulations with no GxE. (a) Proportion of significant tests for genetic correlation across 3 scenarios. (b) Proportion of significant tests for PRSxE regression across 3 scenarios. (c) Proportion of significant tests for SNP-heritability by E across 3 scenarios. Error bars denote standard deviations across 100 simulation replicates.

**Figure S2.**
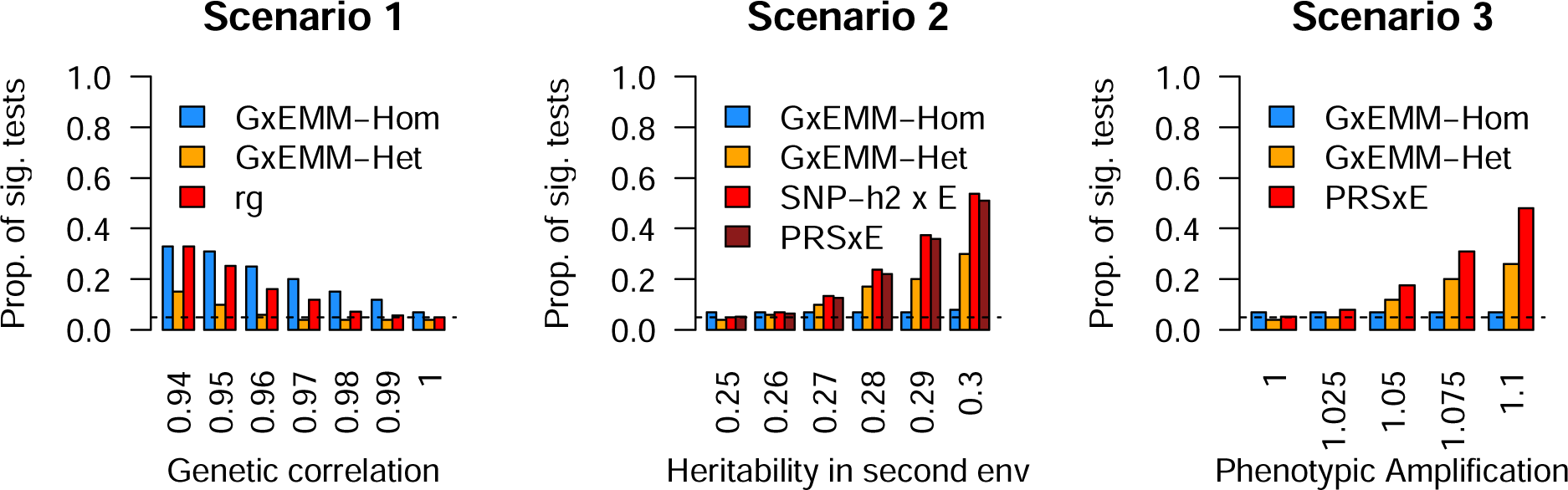
Comparison of three statistical approaches for detecting GxE to ExEMM in simulations. a) Scenario 1 with varying true genetic correlation across E, b) Scenario 2 with varying heritability in the second environment, c) Scenario 3 with phenotypic amplification across E bins.

**Figure S3.**
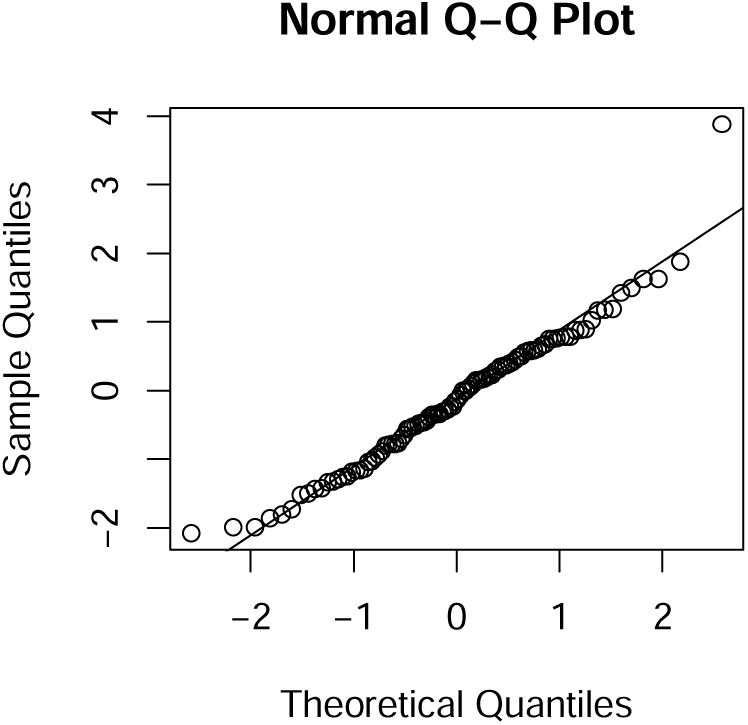
Null simulations of a heritable and genetically correlated E variable. We simulated an E variable that is 100% genetically correlated to the phenotype with the same heritability (h^2^=25%). Each point is the result of a PRSxE regression test from a single simulation (N=100 simulations). We find no inflation of the test statistic under this null model.

**Figure S4.**
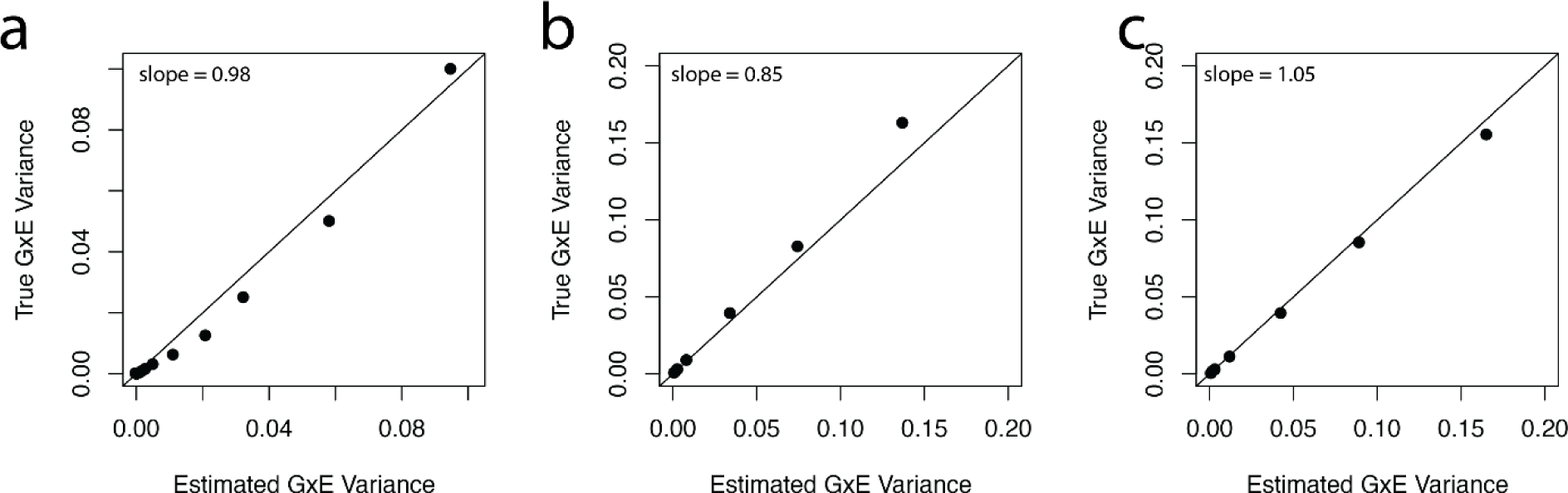
Accuracy of estimates of excess trait variance explained by GxE interaction in simulations. a) genetic correlation in Scenario 1, b) PRSxE in Scenario 2, and c) PRSxE in Scenario 3. For all plots, the black line corresponds to the y=x line and the x and y axes are both on a log scale.

**Figure S5.**
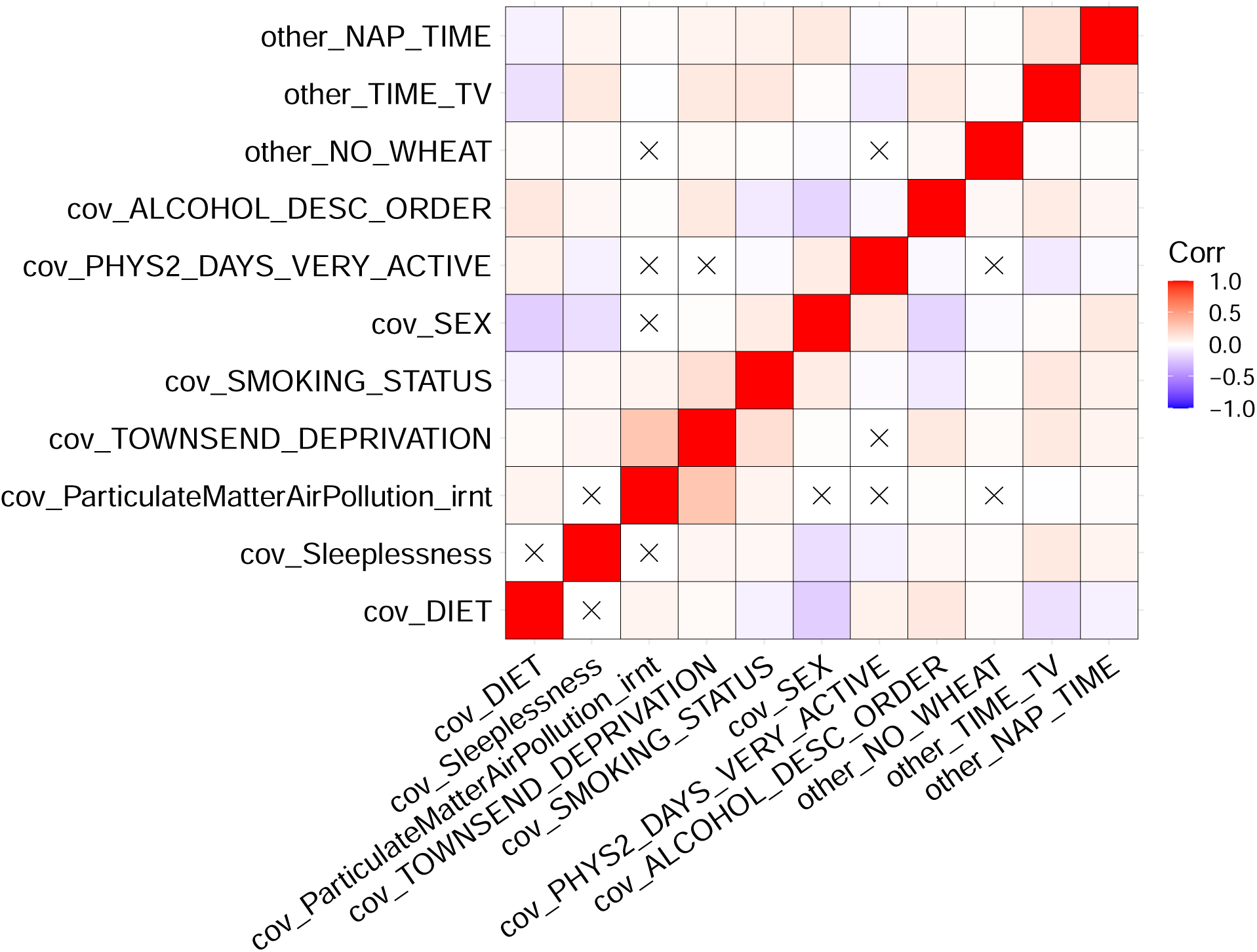
Phenotypic correlations between E variables. X denotes non-significant comparisons at a p-value threshold of 0.05/11.

**Figure S6.**
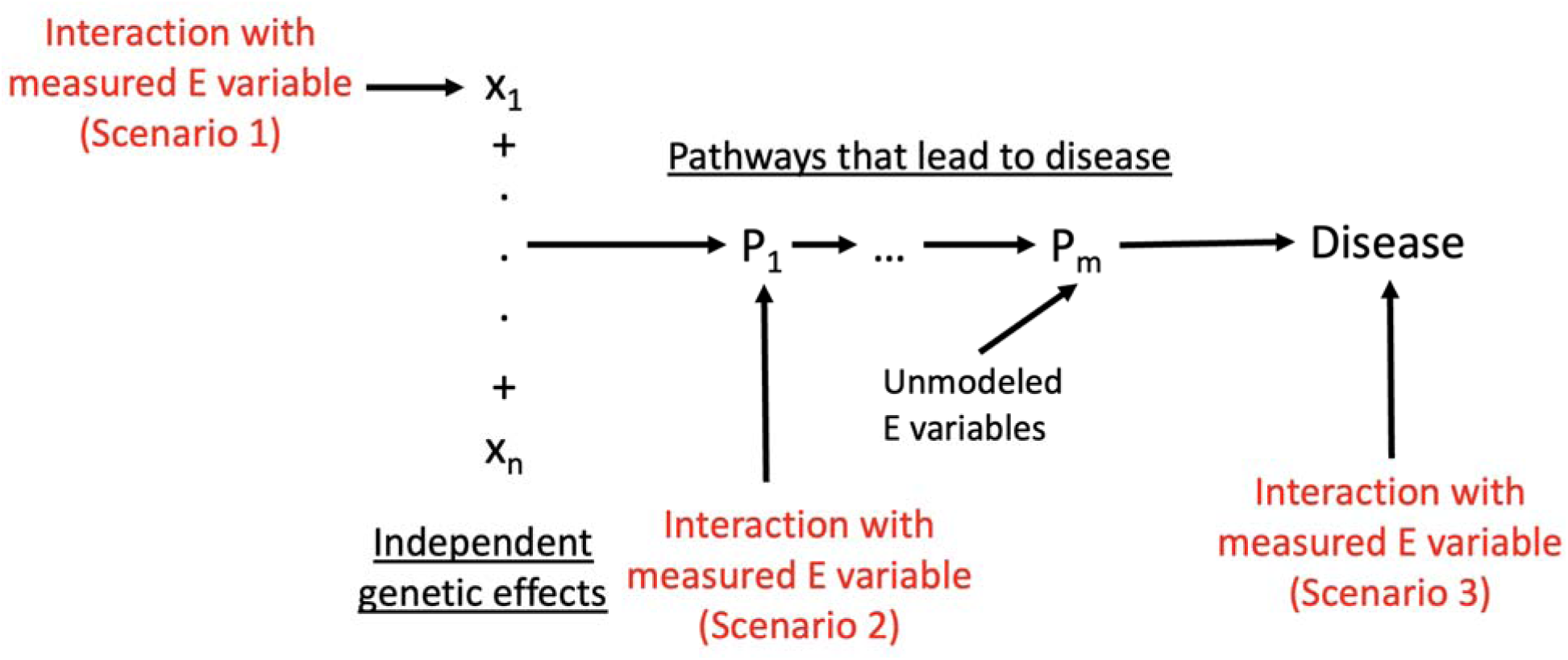
Conceptual model linking three scenarios of GxE. Scenario 1 can be conceptualized as E variables modifying the effects of independent loci. Scenario 2 can be conceptualized as modifying pathways which aggregate the genetic effects of many loci, resulting in a scaling of genetic effects. Finally, Scenario 3 can be conceptualized as modifying the total genetic liability.

**Figure S7.**
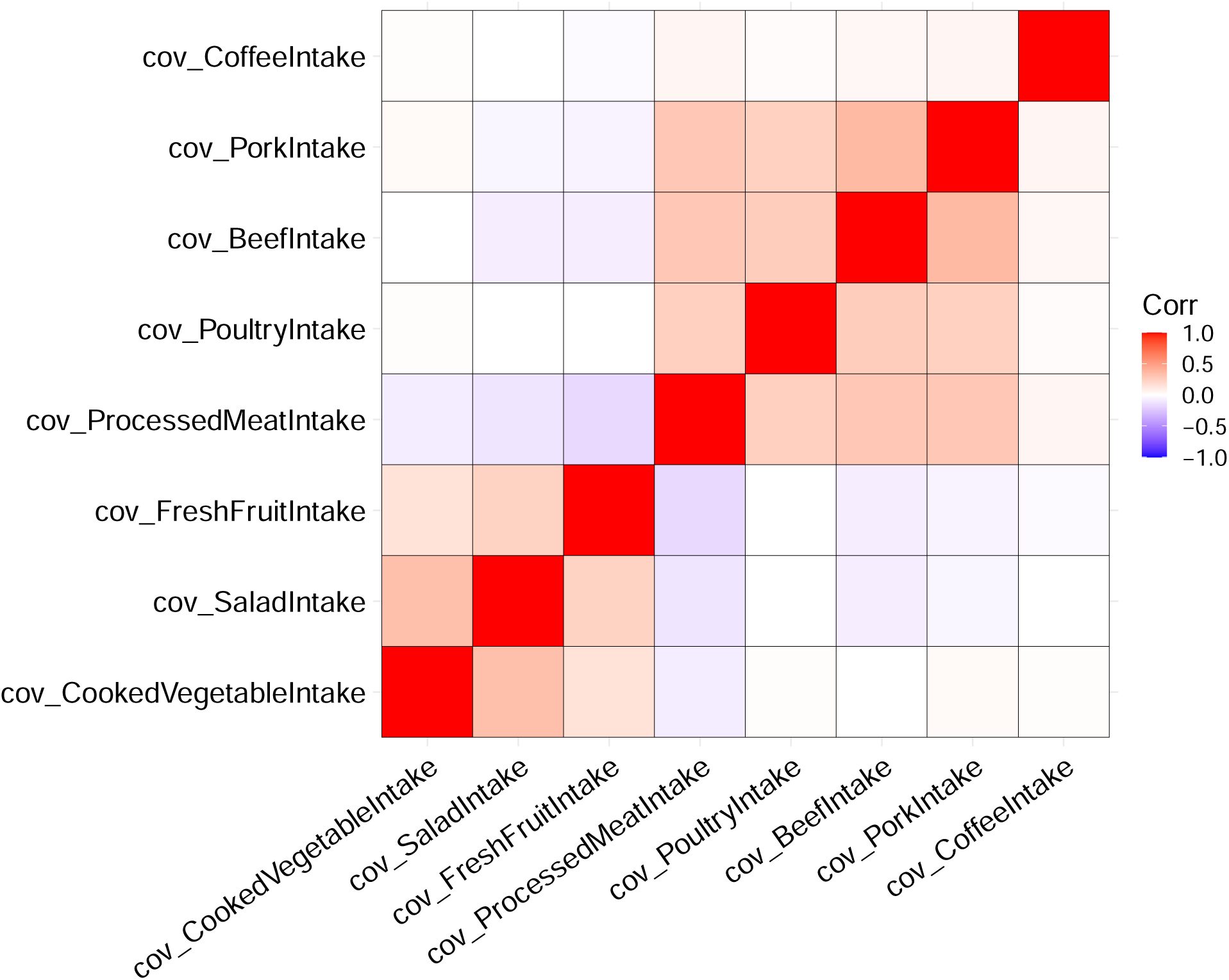
Phenotypic correlations of diet variables used to construct a composite Diet variable. Each cell in the heatmap shows the correlation between the measured diet variable on the X axis and the measured diet variable on the Y axis. All correlations are significant at a Bonferroni corrected p-value threshold of 0.05.

**Supplementary Table 1 Numerical results of Detecting and distinguishing between 3 Scenarios of GxE interaction in simulations.** For each statistical approach and scenario, we report the proportion of significant tests and standard deviation across replicates.

**Supplementary Table 2 Simulations showing bias induced by correlated G and GxE effects.** We tested the impact of correlated G and GxE effects on variance component estimates when assuming that G and GxE effects are not correlated. We set the true variance of G effects to 0.1 and the true variance of GxE effects to 0.1. We varied the correlation of the G and GxE effects and simulated values for 100,000 individuals. We estimate the variance explained by G and GxE using ANOVA in R and report the bias (estimated effect - true effect).

**Supplementary Table 3 Description of the 33 UK Biobank traits analyzed.** For each trait we report a detailed name, the GWAS sample size (including number of cases for binary traits), the SNP-heritability (liability scale for binary traits), and the PRS accuracy (R^2^; observed scale for binary traits).

**Supplementary Table 4 SNP-heritability of E variables studied here.** We estimated SNP-heritability using LDSC. For the composite diet variable, we report the SNP-heritability for each of the underlying variables that make up the composite diet variable. P-values test against a null of zero SNP-heritability.

**Supplementary Table 5 Numerical results of Detecting, quantifying, and distinguishing between 3 Scenarios of GxE interaction across 33 traits and 10 E variables.** For each trait-E pair (A, B), we report (C) the excess variance explained by PRSxE and (D) the associated q value, (E) the difference in heritability between the top and bottom bins of the E variables and (F) the associated q value, (G) the genetic correlation between the top and bottom bin of the E variable and (H) the associated q value. We also assign each trait-E pair to the three scenarios (I, J, K).

**Supplementary Table 6 P-values using robust regression in PRSxE regression analysis compared to P-value from the main PRSxE regression analysis.** For each trait-E pair, we report (A) p-value and (B) effect size for the main PRSxE regression and (C, D) using the Huber-White variance estimator (robust regression).

**Supplementary Table 7 Comparison of disease and quantitative trait variance explained for Scenario 2 GxE.** For 3 matched pairs of disease and quantitative traits, we compared the variance explained on the observed scale for diseases and quantitative trait scale for quantitative traits.

**Supplementary Table 8 SNP-heritability differences for trait-E pairs with no PRSxE interaction**. For each trait-E pair with a significant difference in SNP-heritability and no significant PRSxE interaction we report the SNP-heritability difference and q-value at 5% FDR control.

**Supplementary Table 9 Numerical results of Examples of 3 Scenarios of GxE interaction.** We report detailed results for 3 trait-E pairs reported in Figure 4. For each trait-E pair (A, B), we report (C) the genetic correlation and (D) p-value, (E) PRSxE regression coefficient and (F) p-value, (G, H, I, J, K) SNP-heritability across bins of the E variable with associated standard error and (L) the p-value testing for a difference between the top and bottom bins of the E variable.

**Supplementary Table 10 Numerical results of Detecting, quantifying, and distinguishing between 3 Scenarios of GxSex interaction across 33 traits. For each trait (A, B), we report** (C) the excess variance explained by PRSxSex and (D) the associated q value, (E) the difference in heritability between males and females and (F) the associated q value, (G) the genetic correlation between males and females and (H) the associated q value. We also assign each trait-E pair to the three scenarios (I, J, K).

**Supplementary Table 11 SNP-heritability differences for trait-sex pairs with no PRSxSex interaction.** For each trait-sex pair with a significant difference in SNP-heritability and no significant PRSxSex interaction we report the SNP-heritability difference and q-value at 5% FDR control.

**Supplementary Table 12 Comparison of disease and quantitative trait variance explained for Scenario 2 GxSex.** For 3 matched pairs of disease and quantitative traits, we compared the variance explained on the observed scale for diseases and quantitative trait scale for quantitative traits.

**Supplementary Table 13 Numerical results of Examples of 3 Scenarios of GxSex interaction.** We report detailed results for 3 trait-sex pairs reported in Figure 4. For each trait (A, B), we report (C) the genetic correlation and (D) p-value, (E) PRSxE regression coefficient and (F) p-value, (G, H, I, J, K) SNP-heritability across sex with associated standard error and (L) the p-value testing for a difference between the top and bottom bins of the E variable.

**Supplementary Table 14 Average variance explained by binary and quantitative traits.** For each scenario, we report the average trait variance explained by binary, quantitative, and all traits for both GxE and GxSex interactions.

**Supplementary Table 15 PRSxE regression results including a non-linear E term.** For each trait-E pair, we report (A) P-value and (B) effect size including E^2^ as a covariate and (C, D) not including E^2^ as a covariate.

**Supplementary Table 16 Trait variance explained by GxE interactions with multiple E variables.** For traits with multiple marginally significant E variable interactions, we report the variance explained by a joint model with all marginally significant E variables.

**Supplementary Note for “Distinct explanations underlie gene-environment interactions in the UK Biobank”**

