## Supplementary note for "Distinct explanations underlie gene-environment interactions in the UK Biobank"

April 2024

#### Transformation of genetic correlation across E bins to variance explained by GxE

Consider a population where individuals are exposed to a binary environmental variable and we are modeling individuals with low ($l$) and high ($h$) values. $X$ is an $n \times m$ mean-centered and standardized individual-by-SNP matrix, $\beta$ is a length $m$ vector of environment-specific SNP effect sizes, and $e$ is a length $n$ vector of environmental noise. The SNP effects $\beta$ and environmental noise $e$ are random vectors, while the genotype matrix is fixed. We model the phenotypes $y$ as

$$\begin{aligned} y_l &= X_l \beta_l + e_l \\ y_h &= X_h \beta_h + e_h \end{aligned}$$

$$\begin{aligned} \begin{pmatrix} \beta_l \\ \beta_h \end{pmatrix} &\sim \mathcal{N}\left(\mathbf{0}, \begin{pmatrix} \sigma_g^2 & \gamma \\ \gamma & \sigma_g^2 \end{pmatrix}\right) \\ \begin{pmatrix} e_l \\ e_h \end{pmatrix} &\sim \mathcal{N}\left(\mathbf{0}, \begin{pmatrix} \sigma_e^2 & 0 \\ 0 & \sigma_e^2 \end{pmatrix}\right) \end{aligned}$$

The variance within the low bin population is

$$\mathrm{Var}(y_l) = \sigma_g^2 \frac{X_l X_l^T}{M} + \sigma_e^2 \mathbf{I}_N$$

and the analogous variance for the high bin population is obtained by substituting the corresponding phenotype vector and genotype matrix. The covariance between the low and high phenotypes is

$$\begin{aligned} \mathrm{Cov}(y_l y_h) = \mathbb{E}\left(y_l y_h^T\right) &= \mathbb{E}\left((X_l \beta_l + e_l)(X_h \beta_h + e_h)^T\right) \\ &= \mathbb{E}\left(\beta_l \beta_h^T\right) \frac{X_l X_h^T}{M} \\ &= \gamma \frac{X_l X_h^T}{M} \end{aligned}$$

where we have assumed no covariance between environmental effects and genetic effects between environmental variables and no covariance between environmental effects.

To obtain the variance of the whole population, we average across the bins. Define the mean genetic effects over the low and high bins $\beta = \frac{1}{2}(\beta_l + \beta_h)$. Using the definition of the variance, we see that

$$
\begin{aligned}
\mathrm{Var}(\beta) &= \mathrm{Var}\left(\frac{1}{2}(\beta_l + \beta_h)\right) \\
&= \frac{1}{4}\left(\mathrm{Var}(\beta_l) + \mathrm{Var}(\beta_h) + 2\mathrm{Cov}(\beta_l, \beta_h)\right) \\
&= \frac{1}{4}\left(\mathbb{E}(\beta_l^2) + \mathbb{E}(\beta_h^2) + 2\,\mathbb{E}(\beta_l \beta_h)\right) \\
&= \frac{1}{4}\left(\sigma_g^2 + \sigma_g^2 + 2\gamma\right) \\
&= \frac{\sigma_g^2 + \gamma}{2}
\end{aligned}
$$

In the extreme case of no correlation between the effects ($\gamma = 0$), we see that the genetic variance is halved.

Define the genetic correlation using the standard definition of a bivariate normal correlation $r = \frac{\gamma}{\sigma_g \sigma_g}$. Rearranging $\gamma = r * \sigma_g^2$ implies

$$
\mathrm{Var}(\beta) = \frac{\sigma_g^2(1 + r)}{2}
$$

Then, we can define the following quantities scaled by $\sigma_g^2$:

- Genetic variance $\mathrm{Var}(\beta) = (1 + r)/2$
- Total variance $\mathrm{Var}(y) = 1$
- Total variance minus genetic variance $= 1 - (1 + r)/2 = (1 - r)/2$

The final quantity is the excess variance explained by GxE.

### Expanded description of test outcomes and scenarios

In the Main text, we describe three distinct tests, each of which has a binary outcome. Taken together, there are 8 possible outcomes. Here, we provide interpretations for all outcomes. In the table, 0 indicates a test is not considered significant and 1 indicates that it is considered significant. Two of the seven outcomes result in no GxE, while two of the seven outcomes result in multiple scenarios of GxE.

| Genetic correlation | PRSxE Regression | SNP-heritability by E | Interpretation |
|---|---|---|---|
| 0 | 0 | 0 | No GxE |
| 1 | 0 | 0 | Scenario 1: SNPs are multiplied by SNP-specific factors (locus-dependent GxE) |
| 0 | 1 | 0 | Scenario 3: phenotype is multiplied by an environment-specific vator (non-locus-dependent GxE) |
| 0 | 0 | 1 | Change in environmental variance; No GxE |
| 1 | 1 | 0 | Scenario 1 and 3: SNPs are multiplied by SNP-specific factors and the overall phenotype is also multiplied by factor (locus-dependent and non-locus dependent GxE) |
| 1 | 0 | 1 | Scenario 1 with change in environmental variance: SNPs are multiplied by SNP-specific factors (locus-dependent GxE) and there is a change in the amount of environmental variance contributing to the trait |
| 0 | 1 | 1 | Scenario 2: the genetic variance is multiplied by a factor (non-locus depedent GxE) |
| 1 | 1 | 1 | Scenario 1 and 2: SNPs are multiplied by SNP-specific factors and the genetic variance is also multiplied by a factor (locus-dependent and non-locus dependent GxE) |
